# Targeted Screening of Unruptured Intracranial Aneurysms in 50- to 60-Year-Old Female Smokers – Inconsiderate Overdiagnosis or a Chance to Improve Women’s Health?

**DOI:** 10.1101/2024.12.11.24318889

**Authors:** Justiina Huhtakangas, Ville Vasankari, Jussi Numminen, Mika Niemelä, Miikka Korja

**Affiliations:** Department of Neurosurgery, University of Helsinki and Helsinki University Hospital, P.O. Box 266, FI-00029 Helsinki, Finland; Department of Radiology, University of Helsinki and Helsinki University Hospital, P.O. Box 266, FI-00029 Helsinki, Finland

**Keywords:** Prevention, Screening, Smoking, Unruptured Intracranial Aneurysms, Aneurysmal Subarachnoid Hemorrhage, Womeńs Health

## Abstract

**Introduction:** Aneurysmal subarachnoid hemorrhage (aSAH) causes a substantial proportion of all deaths among middle-aged people, especially women. Since female smokers in particular have a high risk of aSAH and SAH deaths, targeted screening of 50–60-year-old female smokers could be justified as a preventive action to reduce premature deaths and morbidity.

**Methods:** This prospective screening study has been carried out at Helsinki University Hospital in the Department of Neurosurgery in two phases during 2020 and 2023–2024. In order to minimize recruitment bias, the Helsinki Biobank and THL Biobank were responsible for sending out preliminary invitation letters to self-caring 50- to 60-year-old women who were known to be active smokers. We informed the potential candidates about the study and answered any questions before their decision to participate. Once written consent was provided, participants filled in a detailed questionnaire on lifestyle and health, and underwent computed tomography angiography (CTA) analysis. We studied the prevalence of unruptured intracranial aneurysms (UIAs) among the study participants. Moreover, we assessed morbidity, mortality and costs related to screening.

**Results:** Of the 458 preliminary invitation letters, 160 potential participants initially replied. Of these, 116 returned questionnaires and written consents. Ultimately, 108 smoking women underwent CTA imaging. Eleven UIAs were found in eleven (10%) female smokers, one of which was intracavernous and extradural. Two women were operated on without complications – one with a middle cerebral artery aneurysm and one with a posterior communicating artery aneurysm. Most (n=8) patients with small (<5mm) intradural aneurysms were treated with non-invasive preventive actions (smoking cessation, blood pressure control) and followed with CTA or MRA thereafter.

**Conclusions:** Small UIAs, which may be best suited for non-invasive preventive actions, seem to be common in 50- to 60-year-old female smokers. Targeted screening of women in this age group may offer a chance to reduce morbidity or mortality of aSAH. Whether this leads to improved health and reduction in SAH-related deaths in the long run requires a longer follow-up.

## INTRODUCTION

Case-fatality rates (CFRs) of aneurysmal subarachnoid hemorrhage (aSAH) are slowly decreasing (1). Around 25% of SAH patients experience sudden death out of hospital (2,3,4). This risk seems to be highest among individuals with the most adverse risk profile, i.e., hypertensive heavy smokers (2). In line with this, earlier epidemiological studies have shown that female heavy smokers are at the highest risk of aSAH (2,4,5). Among females, the annual risk for aSAH may be as high as 8-fold in smokers compared to never-smokers (2). Furthermore, the burden of aSAH deaths is exceptionally high in middle-aged women (6). This is still the case even though smoking rates as well as the incidence of aSAH (7/100 000 in Finland) are decreasing worldwide (3,7).

Screening programs have been under critical observation during the past years (8–10). In fact, earlier diagnosis does not benefit the patient unless it decreases morbidity or mortality. Since people with unruptured intracranial aneurysms (UIAs, prevalence 2-3% in general (11)) are mainly asymptomatic, and not all aneurysms rupture during their lifetime, searching for UIAs in asymptomatic people may lead to overdiagnosis and overtreatment. Therefore, UIA screening should be restricted to high-risk people, if any.

Nowadays, screening is offered on the basis of family history of intracranial aneurysms (IAs). Moreover, in certain medical conditions, such as autosomal dominant polycystic kidney disease (ADPKD), screening of UIAs is recommended (12). The goal of our screening study is to estimate if targeted screening of people at an exceptionally high risk of aSAH would detect a high prevalence of UIAs. In our previous pilot, we screened 43 women and found five (12%) individuals with UIAs. Based on these positive preliminary results, we received permission to extend the screening to a larger population. To have a realistic view on risks of screening, we also report screening- and treatment-related morbidity, mortality and costs. Our hypothesis is that about one in ten smoking 50- to 60-year-old women has an UIA. If this is accurate, it is worth discussing whether similar targeted screening studies should also be launched in other countries.

## METHODS

### Recruitment protocol

Our research protocol has been published earlier (13). In short, to minimize recruitment bias, the Helsinki Biobank and The Finnish Institute for Health and Welfare (THL) Biobank sent preliminary invitation letters to independent female smokers between the ages of 50 and 60 years old. In 2020, the preliminary invitations were sent by the THL Biobank, which could identify women who had previously replied to a GeneRISK study questionnaire about smoking. In 2023, the preliminary letters were sent by the Helsinki Biobank. These candidates were known to be smokers, as this information was provided in some of their earlier medical reports. These individuals were statistically sieved by a data analyst at the Biobank.

The preliminary invitation letter briefly explained the UIA screening study protocol and rationale. If a person was interested in the screening study and replied to the letter, they were provided full invitation letters, which included a detailed description of the study protocol. All candidates had an opportunity to talk with researchers (JH or MK) if they had further questions or considerations. It was emphasized that participating was voluntary, and that patients had a chance to quit at any stage of the study without justification. All invited participants who returned the signed consent were eligible to self-book a brain CTA examination appointment at the study hospital. Prior to the brain CTA examination, participants were also instructed to have a plasma creatinine test in any of the multiple University Hospital laboratories around Southern Finland. People with kidney failure or iodinated contrast allergy were excluded. Participation in the study was not compensated, but documented travel expenses (use of own car or public transport) were reimbursed.

### CTA screening protocol

The screening protocol has also been published earlier (13). CTA screenings were performed at the Helsinki University Hospital with a multi-slice CT-scanner (GE Lightspeed VCT 64, GE Healthcare, USA) using the standard institutional protocol (120-kV tube potential, NI 10/100-500 mA). A manual bolus tracking was used for the common carotid artery with a contrast material (350 mg I/mL) introduced at an injection rate of 5 mL/s, followed by 40 mL of saline. Axial raw data images with a slice thickness of 0.625 mm were reconstructed. We used 3D workstation servers (syngo.via, Siemens Healthineers; or Vitrea, Vital, Canon Medical Systems), and images were analyzed by neuroradiologists (JN) and neurosurgeons (MK or JH). Any findings with uncertainties were verified with digital subtraction angiography (DSA). Study participants with abnormal imaging findings were invited to the outpatient clinic at the Department of Neurosurgery. Participants without abnormalities were informed by mail. Incidental findings were also reported.

### Number Needed to Screen

The number needed to treat (NNT) is calculated using absolute risk reduction (14), and is represented by the inverse of this reduction. Using the same logic, we calculated the number needed to screen (NNS-opt) with the most optimistic estimation (all screened women quit smoking), as well as the number needed to screen (NNS-pes) with the most pessimistic estimation (no one quits smoking) to be able to theoretically avoid one aSAH or one aSAH death.

## RESULTS

### Study cohort

Of the 108 voluntary participants, one was intersex or non-binary. All had been smoking for decades, ranging from 19 to 50 years (mean 35 yrs, median 37 yrs). Five women reported that they had quit smoking recently (three within one month, two within 1–6 months). Five women smoked every once in a while, and 98 regularly. Participants smoked approximately 12 cigarettes daily (range 0–25, mean 12).

Most women reported their health to be very good (n=5, 5%) or fairly good (n=47, 44%), and nearly as many expressed it as being average (n=43, 40%). Few felt that their health was fairly poor (n=13, 12%). All women had four or more chronic illnesses. Out of all women, 55 (51%) currently or previously had a hypertension diagnosis. Other common medical conditions were hypercholesterolemia (n=39, 36%), asthma or COPD (n=24, 22%), depression (n=13, 12%) and cancer (n=10, 9%).

### Screening results

Of the 458 preliminary invitation letters, 160 potential participants initially replied. Of these, 116 returned questionnaires and signed consents, and 108 underwent CTA analysis. UIAs were found in eleven (10%) women. One UIA was intracavernous and extradural. One patient had a suspected small aneurysm, which turned out to be an infundibular dilation of the posterior communicating artery in DSA.

### Incidental Findings

Three participants had incidental findings. Two had partly calcified and asymptomatic meningeomas, one of which had been diagnosed earlier. Another participant had a dural arterio-venous fistula (dAVF) at the skull base, which was treated surgically without complications.

### Treatment and Follow-Up

Most women with UIAs (n=8) were treated conservatively. Specifically, they were encouraged to quit smoking to minimize the risk of aneurysm growth and rupture, as well as adviced to follow blood pressure levels if not done recently. Follow-up with MRA or CTA was planned for all of these women at one year after the diagnosis.

Two patients were surgically treated for their UIAs. For the first one, neurosurgical clipping was recommended for the 7mm medial cerebral artery (MCA) aneurysm. The surgery was uneventful. For the other patient with a small 2mm posterior communicating (PComA) aneurysm, conservative treatment was strongly recommended. After comprehensive discussions on all treatment options including conservative, endovascular and microsurgical treatments, the patient opted to undergo surgery due to psychological stress related to the diagnosed UIA and their inability to quit smoking. The operated patients were discharged at day two and day four after the operation, respectively, and they both returned to their normal activities without any neurological deficits or other complications within four weeks following surgery. For one treated woman, return to work was delayed due to anxiety following UIA diagnosis and treatment, potentially related not only to the UIA but also to mental disorders.

### Number Needed to Screen

Table 1 shows the results of screening and the proportions of women who ended up in the active or conservative treatment groups. Based on these numbers, an evaluation was made for screening and treatment of 1000 patients and the number of lifetime aSAHs or premature deaths potentially avoided in cases when everyone quits smoking (the most optimistic evaluation) and when no one quits smoking (the most pessimistic evaluation). As a comparison, there is a group of 1000 smoking women with no screening (the natural history group). By using the risk reduction, the number needed to screen was calculated: NNS=1/ARR (absolute risk reduction) = 1/(EER-CER), where EER = expected event rate in natural history, and CER = event rate in calculation group.

**Table 1.**
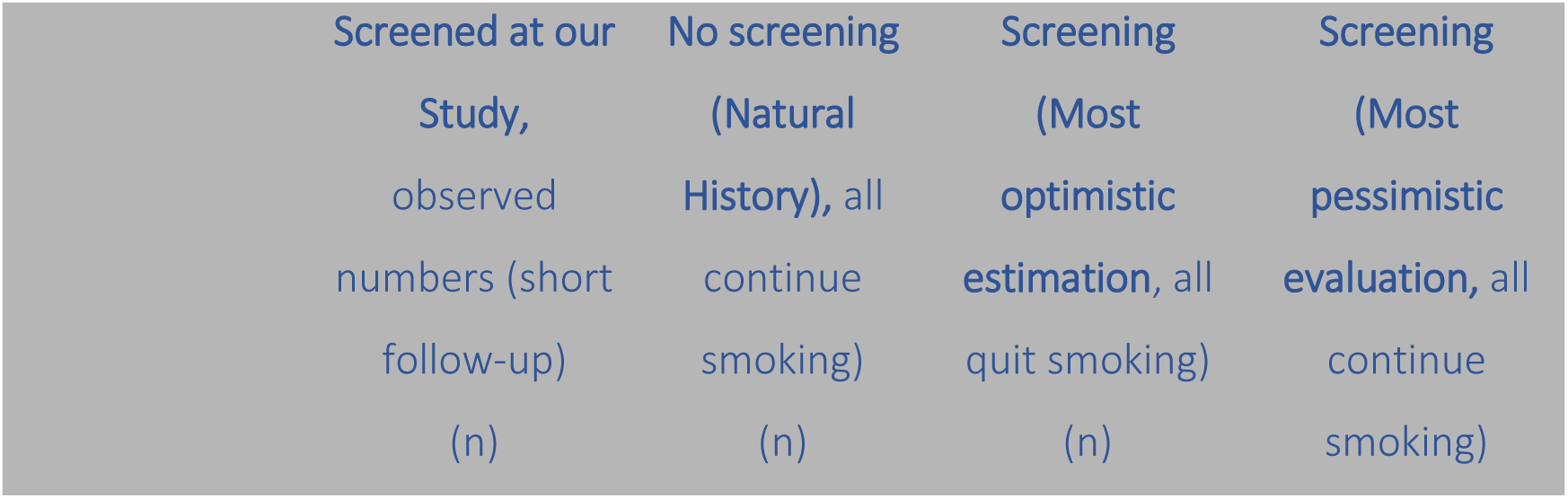
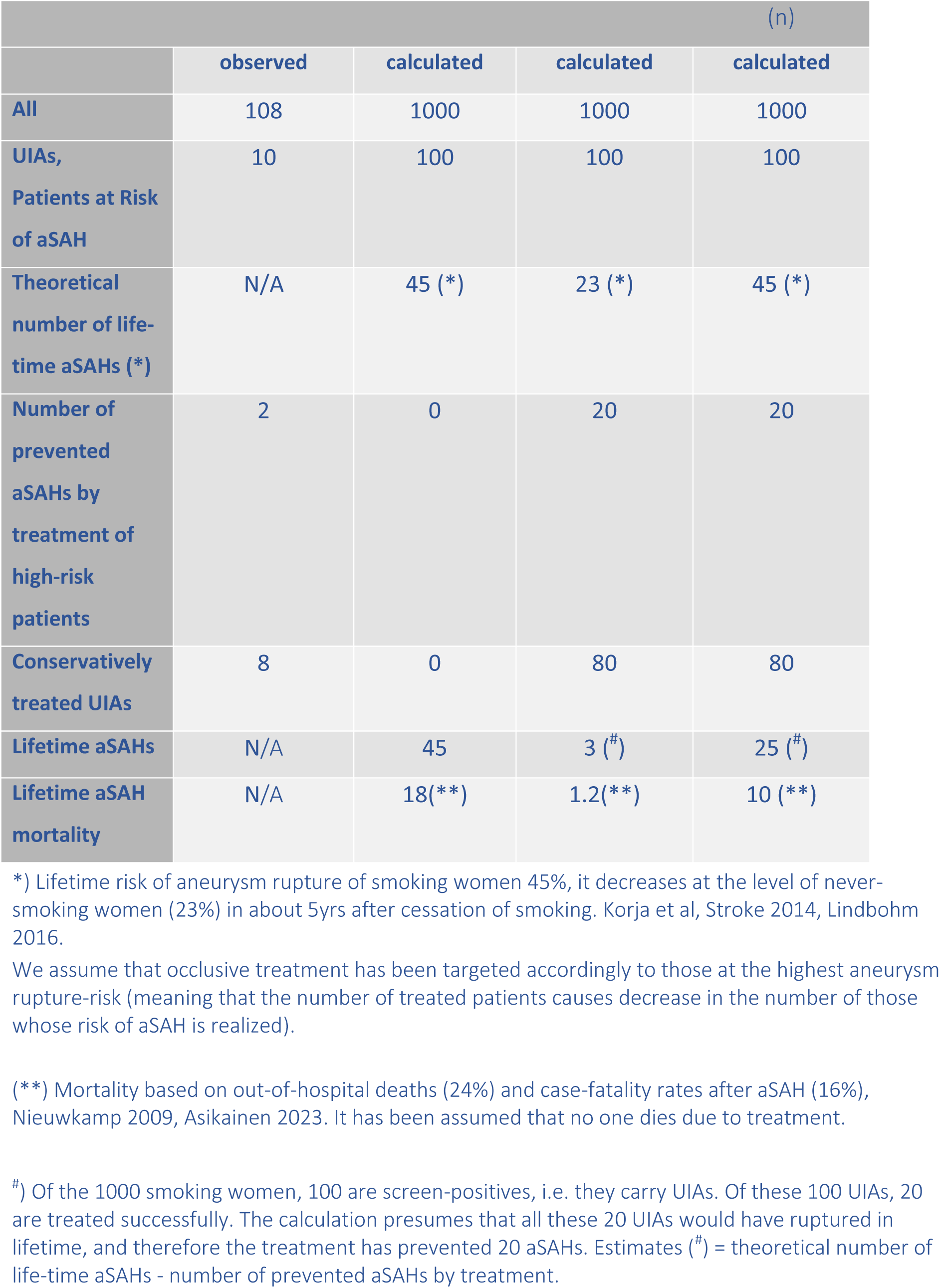
Treatment groups of screened patients, first our detected results (in short-term follow-up), then evaluation on 1000 smoking women based on natural history and screening with two different estimation. We evaluated separately the most optimistic evaluation (all screened individuals stop smoking which leads to decrease in risk of rupture for those treated conservatively) and the most pessimistic evaluation (no screened individual quits smoking when the risk of aneurysm rupture stays at the level of smoking women, which means failure in conservative treatment).

NNS-opt (the most optimistic evaluation) to prevent one aSAH 1/(45/1000-3/1000)=24, NNS-opt to prevent one aSAH death 1/(18/1000-1.2/1000)=60. NNS-pes (the most pessimistic evaluation) to prevent one aSAH 1/(45/1000-25/1000)=50, NNS-pes to prevent one aSAH death 1/(18/1000-10/1000)=125.

### Costs

The total costs of screening and active preventive treatment (microsurgical treatment) of one screen-positive UIA patient were 48 605 EUR (58 440 USD; including screening, treatment and economic costs) in 2020 in the pilot of 43 patients (13). Presuming that the screening costs remained the same in this study, the total costs were about 122 078 EUR for 108 screened individuals (an average of 1,100 EUR per screened individual). By using this amount, the cost to prevent one aSAH (NNS=24-50) would be between 26 400 EUR and 55 00 EUR. The cost to prevent one premature death (NNS=60-125) would be between 60 000 EUR and 137 000 EUR in Finland.

## DISCUSSION

Our results suggest that even one out of ten smoking women in their fifties may carry an UIA. Approximately 11% of middle-aged women in Finland still smoke, which corresponds to about 38,000 women aged 50–60 years old in 2024 (15). In the United States, the proportion of smokers among individuals aged 45 to 65 was 15% in 2021 (16). While some smokers may have health issues, such as pulmonary disease or cancer, this should not preclude efforts to prevent them from developing aSAH, a condition with high mortality rates. Many of these women may still lack information about the risk of developing intracranial aneurysms related to smoking.

There has been great effort to decrease in-hospital mortality related to aSAH. It should not be forgotten that aSAH patients who experience sudden death prior to hospital admission account for around 25% of all aSAH cases (1,3). In Finland, this group of middle-aged women is more substantial than those who die from traffic accidents or brain infarctions in the same age group (6). Moreover, about 16% of hospitalized middle-aged aSAH patients die during the first 30 days due to severe initial bleeding, despite treatment efforts. Viable strategies to reduce the mortality of these women include smoking cessation support, and screening and treatment of UIAs prior to rupture.

There are no data regarding indirect costs resulting from premature death after aSAH. Previous evaluations in Finland have estimated the economic value of premature death due to car accidents (median age of 55, (15)) to be approximately 2 560 000 EUR (17). This estimation includes costs related to emergency response, loss of life, and the monetary valuation of health and productivity. The screening of all 38 000 50- to 60-year-old smoking women in Finland would cost 41.8 million EUR. If the economic value of a premature death (median age of 55 years) was 2.56 million EUR, prevention of only 17 premature deaths in the screened group of 38 000 smoking women would be cost-effective, presuming that screening does not cause morbidity and mortality. Some cost estimates can also be calculated based on years of life lost (YLLs). A value of 23 800 EUR on a life year can be applied to the total life years lost. This value is based on a willingness-to-pay (WTP) estimate of 20 000 GBP (23 800 EUR) for a disease to prevent a life year lost (i.e., NICE threshold value for a cost-effective intervention (18)). This would give a value of 238 000 EUR of indirect cost for every 10 years lost. Based on this estimation, a screening program that would prevent 1760 years of life lost would be cost effective. If presuming that 50- to 60-year-old smoking women have a median of 15 years life to live, a screening program of these smoking women should prevent 117 premature deaths. It should be noted that none of these estimates account for costs related to multiple follow-up CTAs/MRAs and visits.

Historically, research on women’s health has been underfunded and neglected (19). In vascular diseases, only one screening program exists: abdominal aortic aneurysm (AAA) screening for older smoking men aged 65 to 75 (20). The number needed to screen (NNS) to prevent one AAA-related death over ten years ranges from 209 to 769 individuals (21).

Screening of AAAs has been evaluated to be cost-efficient (20,22,23). If our findings can be validated in larger multinational cohorts, and the proportion of unruptured intracranial aneurysms among female smokers remains close to 10%, the NNS would only be 24–50 to prevent one aSAH and 60–125 to prevent one premature death (depending on if the screened individuals are able to quit smoking or not). Furthermore, the premature deaths potentially prevented would occur at younger ages than in smoking men with AAAs. In the meta-analysis of Ying et al., NNS has been calculated using observed reduction in mortality. In our study, this estimation is still based on a theoretical calculation of risk reduction.

The benefits and drawbacks of UIA screening must be thoroughly discussed before proceeding. Consideration should be given to the psychological impact and anxiety that can accompany screening and diagnosis. UIA diagnosis might also affect willingness or possibilities to work or health insurance options. If an UIA is diagnosed but treated conservatively, patient adherence to follow-up and preventive measures is crucial. With proper counseling, neurosurgeons or neurologists can help in alleviating concerns. For very small aneurysms, the rupture risk should be presented realistically, without exaggeration. The optimal follow-up interval and the impact of screening on quality of life remain debated (24).

Our screening study has a few shortcomings. First, we used CTA for screening despite its radiation exposure. However, the radiation exposure from modern CTA is quite low, with a previous study indicating only 0.0011% excess lifetime cancer risk for women undergoing one CTA, and a risk of about 0.0026% after yearly CTAs in a long-term (30–50 year) follow-up (25,26). Furthermore, CTA has a better availability and high specificity (low false positive rate, limited number of incidental findings) when compared to MRA (27). Second, the patient cohort remains relatively small. Therefore, validation using international cohorts is needed to provide crucial information to determine whether expanding screening efforts for smoking women aged 50–60 years old would be advantageous. Third, as smoking rates decline, the incidence of SAH may decrease further, potentially mitigating the suggested benefits of screening. In the long term, systematically addressing risk factors may prove more effective and beneficial than screening and related intervention alone but would require additional health care resources and multi-disciplinary support. Fourth, we did not evaluate the impact of screening on mortality, health-related quality of life and smoking cessation in this study. Therefore, we do not know if the targeted screening could also be an effective way to encourage quitting smoking. If several of the screened individuals succeed in quitting smoking, screening is likely to reduce not only aSAH-related mortality in the long run, but also achieve other health benefits. However, these additional benefits may be challenging to document in detail.

## CONCLUSIONS

Small UIAs may be relatively common (10%) in 50- to 60-year-old female smokers. Targeted screening of this age group may offer a chance to prevent premature death related to aSAH with low-risk methods and low costs, but whether this leads to improved health and reduction of mortality in the long run requires further investigation.

## Data Availability

Pseudonymized research data are personal data according to Finnish legislation and cannot be shared outside the Helsinki University Hospital.

## ACKNOWLEDGEMENTS

We thank the THL Biobank (www.thl.fi/biobank) and Helsinki Biobank for the fluent collaboration in recruiting the study participants and biobank donors for their generous participation in the study.

We would like to thank neurosurgeon Rahul Raj and data analysist Onni Järvinen for statistical considerations and advice related to this study, as well as research nurse Maarit Tuomisto for co-operation and flexibility. Thanks also to Jacquelin DeFaveri for language revision of the manuscript.

## CONTRIBUTORS

MK designed the study and provided guidance with data analysis and interpretation. JH and VV contributed to study design. MK and JH organised and managed the study. MK and JH were responsible for the clinical patient care. JN, MK and JH analysed CTA images. JN performed DSA studies. JH performed the data interpretation. JH and MK drafted the manuscript. All authors critically reviewed and revised the manuscript.

## FIGURE LEGENDS

**Figure 1.**
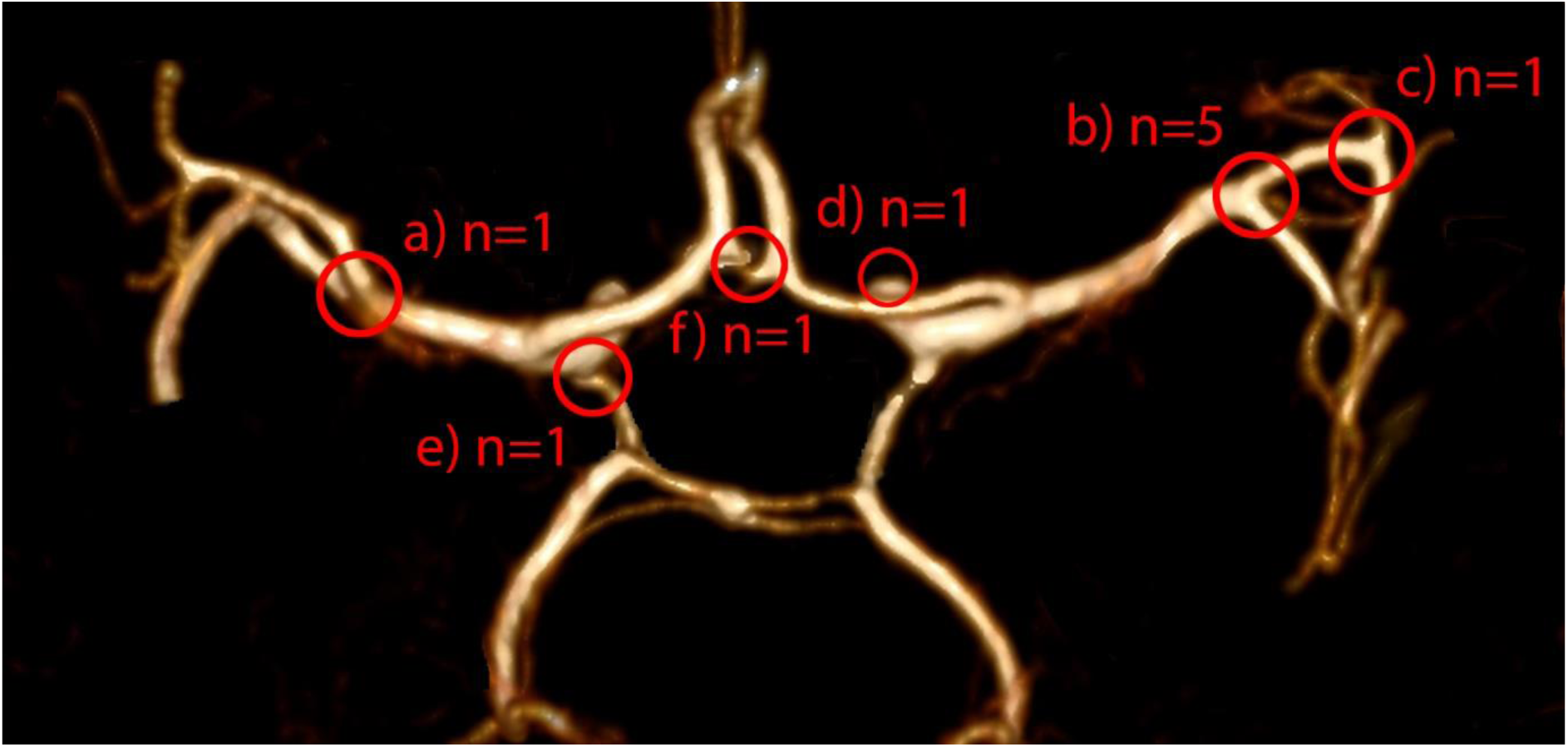
Locations of detected intradural UIAs (n=10) diagnosed as a result of screening: a) middle cerebral artery (MCA) bifurcation on the left side, b) MCA bifurcation on the right, c) M2 branch of MCA on the right, d) internal carotid artery (ICA) - ophthalmic segment on the right, e) ICA -posterior communicating artery segment on the left, f) anterior communicating artery (AComA) in the midline

**Figure 2.**
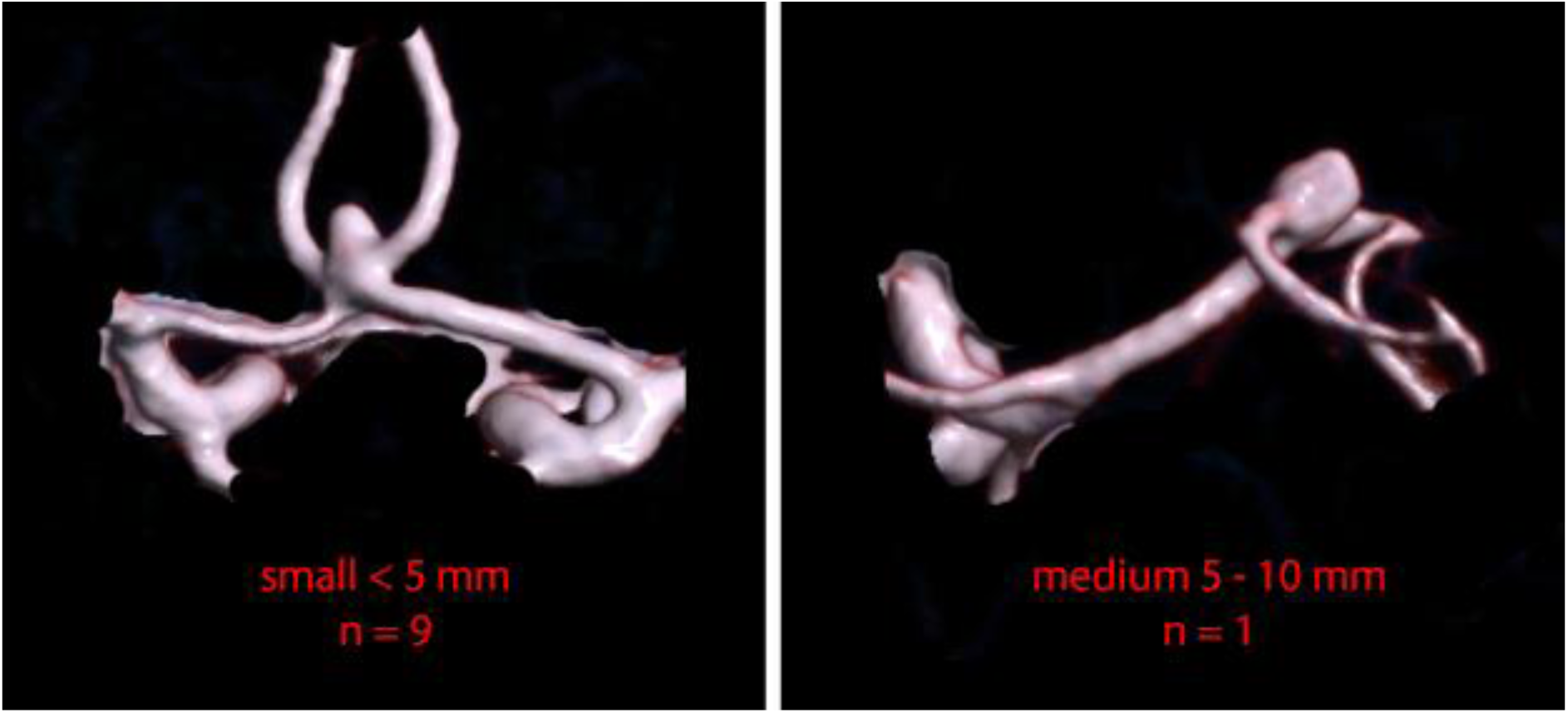
Sizes of detected intradural UIAs (n=10) diagnosed as a result of screening. Most (n=9) were small, meaning the largest dome size being less than 5mm.

